# Glycolysis-enhancing α_1_-adrenergic antagonists modify cognitive symptoms related to Parkinson’s disease

**DOI:** 10.1101/2022.07.01.22277111

**Authors:** Matthew A. Weber, Kartik Sivakumar, Ervina E. Tabakovic, Mayu Oya, Georgina M. Aldridge, Qiang Zhang, Jacob E. Simmering, Nandakumar S. Narayanan

## Abstract

Terazosin is an α_1_-adrenergic receptor antagonist that enhances glycolysis and increases cellular ATP by binding to the enzyme phosphoglycerate kinase 1 (PGK1). Recent work has shown that terazosin is protective against motor dysfunction in rodent models of Parkinson’s disease (PD) and is associated with slowed motor symptom progression in PD patients. However, PD is also characterized by profound cognitive symptoms. We tested the hypothesis that terazosin protects against cognitive symptoms associated with PD. We report two main results. First, in rodents with ventral tegmental area (VTA) dopamine depletion modeling aspects of PD-related cognitive dysfunction, we found that terazosin preserved cognitive function and produced a non-statistically significant trend towards protected VTA tyrosine hydroxylase levels. Second, we found that after matching for demographics, comorbidities, and disease duration, PD patients newly started on terazosin, alfuzosin, or doxazosin had a lower hazard of being diagnosed with dementia compared to tamsulosin, an α_1_-adrenergic receptor antagonist that does not enhance glycolysis. Together, these findings suggest that in addition to slowing motor symptom progression, glycolysis-enhancing drugs protect against cognitive symptoms of PD.

## INTRODUCTION

Parkinson’s disease (PD) is a devastating neurodegenerative disease with motor and cognitive symptoms (Dauer & Przedborski, 2003). A key risk factor for PD is impaired energy metabolism (Saxena, 2012; Wellstead & Cloutier, 2011). We have recently found that the glycolysis-enhancing drug terazosin increases energy metabolism in rodent models and in PD patients (Cai et al., 2019; Schultz et al., 2022). We have also found that terazosin protects against motor neurodegeneration in the MPTP (1-methyl-4-phenyl-1,2,3,6-tetrahydropyridine) mouse model, the 6-hydroxydopamine (6-OHDA) rat model, as well as synuclein-overexpressing mice (Cai et al., 2019). Strikingly, analysis of the Progression Markers Initiative (PPMI) database shows that terazosin is also associated with slowed motor symptom progression in human PD patients (Cai et al., 2019). Furthermore, based on data from large administrative databases, it protects against developing PD (Sasane et al., 2021; Simmering et al., 2021). Notably, patients taking terazosin had fewer cognitive complications, as indexed by International Classification of Diseases (ICD) codes (Cai et al., 2019). Despite these data, it is unclear whether terazosin is neuroprotective for cognitive manifestations of PD.

One challenge is that there are few rodent models of cognitive dysfunction in PD. Human cognition is complex and spans multiple behavioral repertoires such as planning and reasoning, which are challenging to model in rodents. We have found that PD patients with cognitive dysfunction have increased variability during interval timing, which involves estimating an interval of several seconds (Parker et al., 2015; Singh et al., 2021). Specifically, PD patients with cognitive dysfunction have higher interval timing variability compared to PD patients with preserved cognitive function. Because interval timing can be readily studied in rodents (Buhusi & Meck, 2005), and work from our group has demonstrated that interval timing is reliably impaired with disrupted ventral tegmental area (VTA) dopamine (Kim et al., 2017; Kim & Narayanan, 2018; Narayanan et al., 2012; Parker et al., 2015), interval timing provides an opportunity to investigate cognitive deficits in PD rodent models as a function of terazosin.

In this study, we tested the hypothesis that terazosin protects against cognitive symptoms associated with PD. We found that rodents with VTA dopamine depletion via 6-OHDA injections had higher interval timing variability, similar to PD patients with cognitive dysfunction. Subsequently, we found that mice receiving daily terazosin delivered in drinking water had decreased interval timing variability and marginally more tyrosine hydroxylase-positive (TH+) immunofluorescence in the VTA. Since terazosin is prescribed to human PD patients, we were able to combine these results with an investigation of the association between clinically prescribed terazosin and cognitive symptoms in PD in the IBM Marketscan administrative database. We found that human PD patients taking terazosin or related glycolysis-enhancing medications had less risk of developing dementia compared to propensity-score-matched patients taking tamsulosin, an α_1_-adrenergic receptor antagonist that does not enhance glycolysis. Together, these data provide evidence that terazosin protects against cognitive symptoms, as well as motor symptoms, associated with PD.

## RESULTS

### VTA 6-OHDA increases interval timing variability

We trained 29 mice starting at ∼14–16 weeks of age to perform an interval timing task in which mice switched from one nosepoke to another based on internal timing cues to get a food reward (Fig. 1A; Balci et al., 2008; Bruce et al., 2021; Larson et al., 2022). In these animals, we surgically injected vehicle (0.03% ascorbic acid) as a surgical control or 6-OHDA to deplete dopamine in the VTA, which we have previously shown impairs interval timing (Kim et al., 2017; Narayanan et al., 2012; Parker et al., 2015). We began treatment with terazosin or vehicle (dimethyl sulfoxide; DMSO) immediately and tested performance in the interval timing switch task approximately 16 days post-surgery. We normalized post-surgical behavior in the switch task to pre-surgery behavior to determine magnitude change in behavior as a result of VTA dopamine depletion and terazosin treatment. In VTA 6-OHDA mice treated with vehicle, we found no significant change in median switch time (Vehicle DMSO: 103% (93%–117%; relative to presurgical baseline) vs 6-OHDA DMSO: 92% (83%-105%) ; Wilcoxon *p* = 0.12; Cohen’s *d* = 0.66), but increased timing variability as measured by the coefficient of variability (CV) relative to Vehicle DMSO controls (Vehicle DMSO: 93% (86%–104%) vs 6-OHDA DMSO: 118% (116%–123%); Wilcoxon *p* < 0.05; Cohen’s *d* = 1.66; Fig. 2B–C). There were no changes in the number of switch trials performed (Vehicle DMSO: 30 (18–44) vs 6-OHDA DMSO: 38 (30–45); Wilcoxon *p* = 0.60; Cohen’s *d* = 0.23, data not shown) or total number of rewards obtained during short and switch trials (Vehicle DMSO: 102 (95– 124) vs 6-OHDA DMSO: 106 (88–155); Wilcoxon *p* = 0.96; Cohen’s *d* = 0.27, data not shown). These results were similar to the increased interval timing CV observed in PD patients associated with cognitive dysfunction (Singh et al., 2020) and provide evidence that VTA dopamine depletion models aspects of cognitive dysfunction in human PD patients.

**Figure 1:**
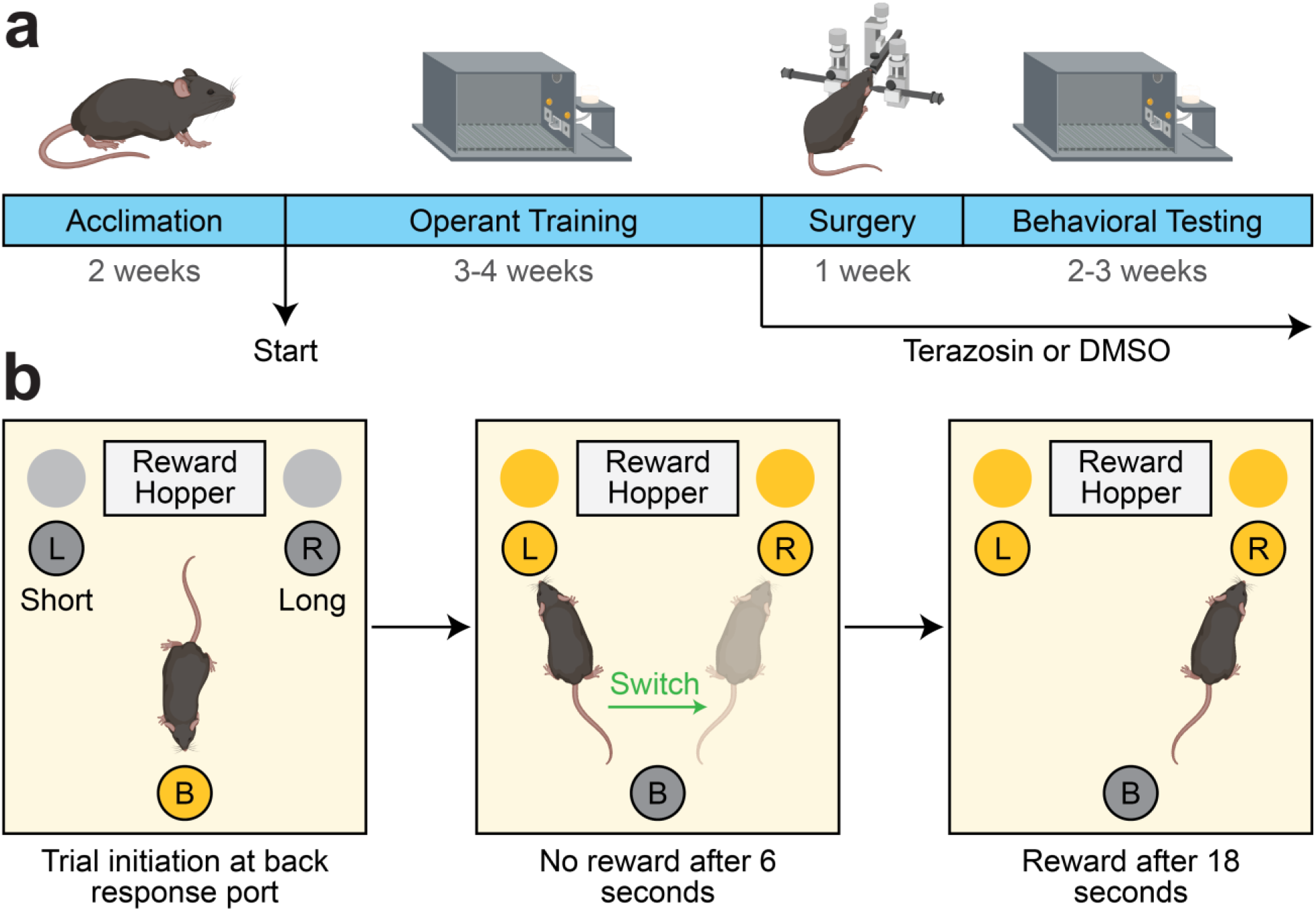
Experimental design. **a**) Experimental timeline. DMSO: dimethyl sulfoxide. **b**) Interval timing switch task highlighting optimal performance during long trials. Trials are initiated at the back response port. Identical cues are delivered for both short and long trials, which are randomly delivered. On short trials, mice are rewarded for the first response after 6 seconds at the designated short nosepoke (left or right). On long trials, mice start by responding at the designated short nosepoke. When there is no reward after 6 seconds, mice switch to the designated long nosepoke (contralateral to designated short nosepoke) and wait 18 seconds for reward delivery. This time to switch from the short to long nosepoke is a time-based decision as in other interval timing tasks. Switch time is defined as the time of last response at the short nosepoke before responses start at the long nosepoke, and only switch trials are analyzed.

**Figure 2:**
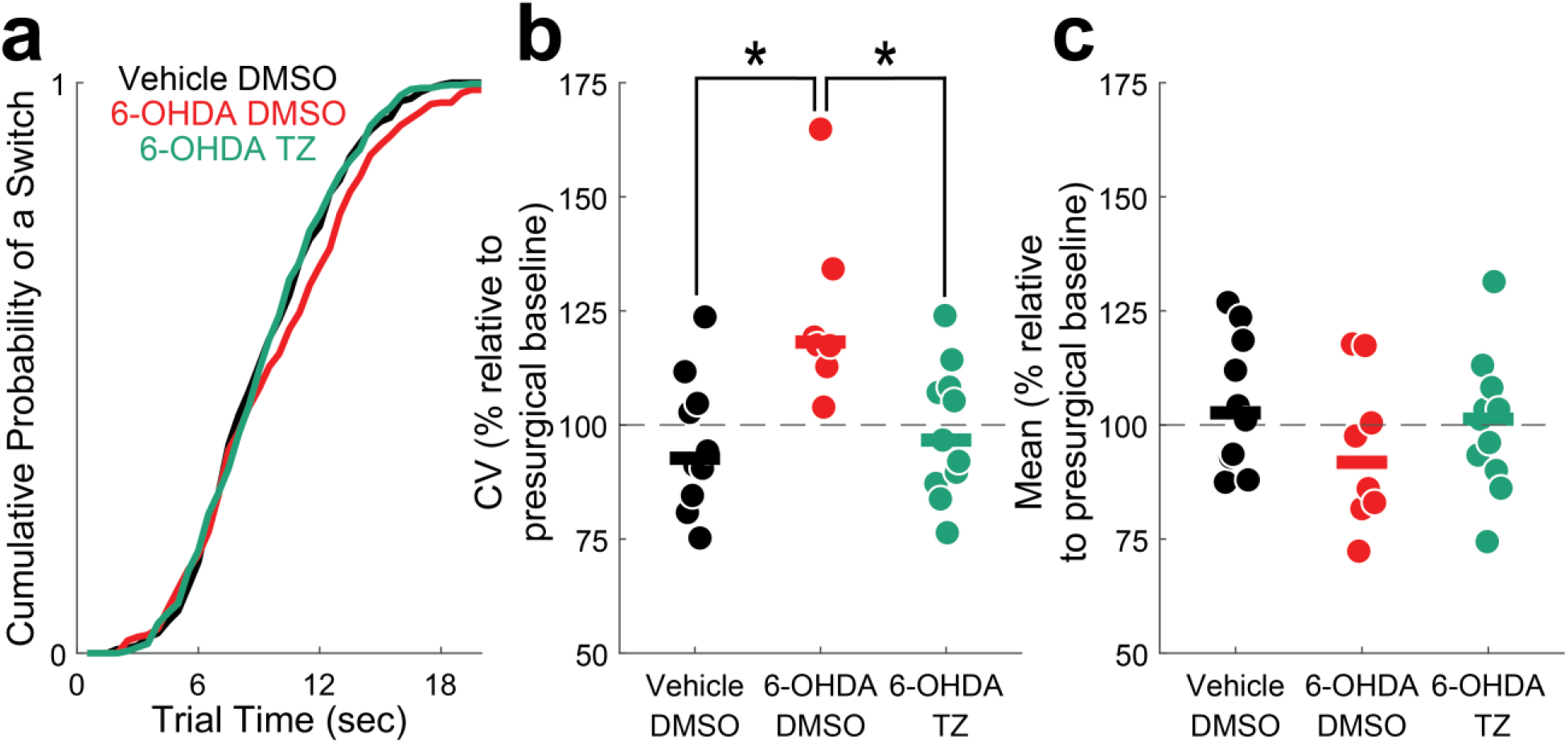
Interval timing variability improves in VTA dopamine-depleted mice treated with terazosin. **a**) Cumulative distribution function of switch times. **b**) Switch time coefficient of variability (CV) and **c**) mean switch time. Data from 10 VTA Vehicle mice treated with dimethyl sulfoxide (DMSO; in black), 8 VTA 6-hydroxydopamine (6-OHDA) mice treated with DMSO (in red), and 11 VTA 6-OHDA mice treated with terazosin (TZ; in green). Each dot represents a single mouse, and the horizontal line represents the median value. All data presented are approximately 16 days post-surgery and normalized according to pre-surgical behavior. * *p* < 0.05.

### Terazosin protects against VTA 6-OHDA timing deficits

Terazosin is neuroprotective in preclinical rodent models of PD-related motor dysfunction (Cai et al., 2019). We investigated whether terazosin protects against the effects of VTA 6-OHDA in interval timing. Strikingly, we found that VTA dopamine-depleted mice treated with terazosin had decreased interval timing variability relative to VTA 6-OHDA mice treated with vehicle (6-OHDA DMSO: 118% (116%–123%) vs 6-OHDA terazosin: 97% (88%–108%); Wilcoxon *p* < 0.05; Cohen’s *d* = 1.53), but no change in median switch time (6-OHDA DMSO: 92% (83%–105%) vs 6-OHDA terazosin: 101% (92%– 106%); Wilcoxon *p* = 0.40; Cohen’s *d* = 0.35; Fig. 2a–c). There were no changes in the number of switch trials performed (6-OHDA DMSO: 38 (30–45) vs 6-OHDA terazosin: 25 (16–51); Wilcoxon *p* = 0.68; Cohen’s *d* = 0.09, data not shown) or total number of rewards obtained during short and switch trials (6-OHDA DMSO: 106 (88–155) vs 6-OHDA terazosin: 113 (97–128); Wilcoxon *p* = 1.0; Cohen’s *d* = 0.07, data not shown). These data support our hypothesis that terazosin is protective in rodents modeling aspects of cognitive dysfunction in PD.

We also examined the levels of TH+ immunofluorescence in the VTA as an indirect measure of dopamine depletion (Fig. 3a), since terazosin has been shown to protect substantia nigra TH levels in rats (Cai et al., 2019) VTA 6-OHDA mice had markedly decreased TH+ fluorescence in the VTA (Vehicle DMSO: 1776 (1577–2010) arbitrary units (AU) vs 6-OHDA DMSO: 673 (623–779) AU; Wilcoxon *p* < 0.05; Cohen’s *d* = 3.07; Fig. 3b). Interestingly, we found a non-statistically significant trend for increased TH+ fluorescence in the VTA of dopamine-depleted mice treated with terazosin relative to DMSO (6-OHDA DMSO: 673 (623–779) vs 6-OHDA terazosin: 1071 (653–1094); Wilcoxon *p* = 0.09; Cohen’s *d* = 0.79; Fig. 3b), suggesting that terazosin tended to protect TH+ neurons in the VTA of dopamine-depleted mice. This effect of terazosin is similar to that observed previously (Cai et al., 2019). To establish a relationship between the change in VTA TH+ immunofluorescence and behavioral results, we correlated fluorescence values with changes in switch time CV following surgery and DMSO or terazosin treatment. We observed that VTA TH+ fluorescence in all DMSO-treated mice had a strong and significant negative correlation with changes in switch time CV, such that greater VTA TH+ fluorescence values were related to lower switch time CV (*r* = -0.59; *p* < 0.05); there was also a similar non-significant relationship between switch time CV and VTA TH+ fluorescence values in dopamine-depleted mice treated with terazosin (*r* = -0.47; *p* = 0.15; Fig. 3c).

**Figure 3:**
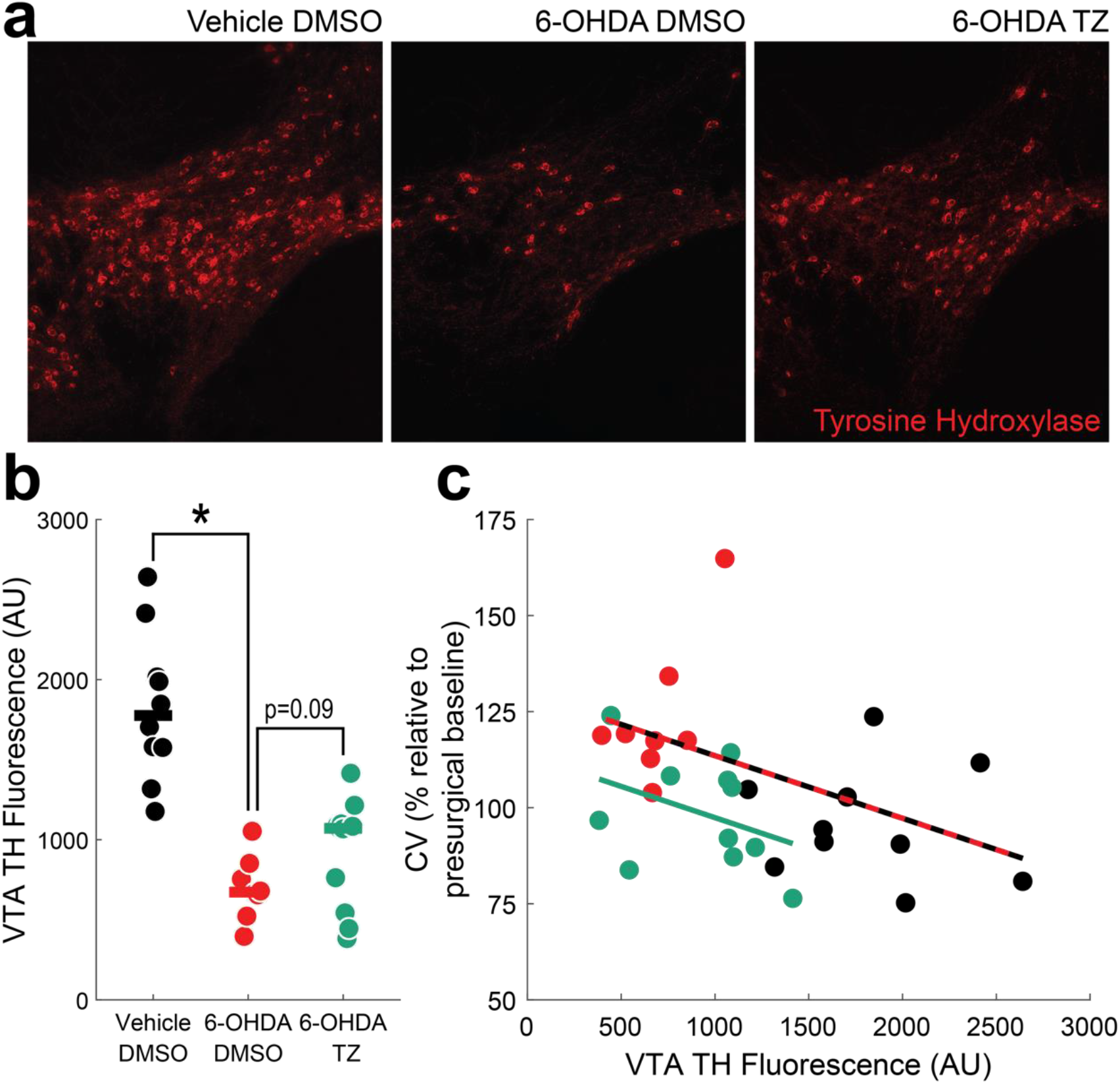
VTA tyrosine hydroxylase (TH) fluorescence levels correlate with interval timing coefficient of variability. **a**) Representative histological images of TH fluorescence (red) from VTA Vehicle mice treated with dimethyl sulfoxide (DMSO; left), VTA 6-hydroxydopamine (6-OHDA) mice treated with DMSO (middle), and VTA 6-OHDA mice treated with terazosin (TZ; right). **b**) VTA TH fluorescence (in arbitrary units; AU) from 10 VTA Vehicle mice treated with DMSO (in black), 8 VTA 6-OHDA mice treated with DMSO (in red), and 11 VTA 6-OHDA mice treated with terazosin (in green). * *p* < 0.05. Each dot represents a single mouse, and the horizontal line represents the median value. **c**) In VTA Vehicle and VTA 6-OHDA mice treated with DMSO, the percent change in coefficient of variability (CV) relative to presurgical baseline is strongly correlated with VTA TH fluorescence (red and black line). The relationship between VTA 6-OHDA and mice treated with terazosin is shown in green.

### TZ protects against cognitive symptoms of PD

An analysis of patient databases was used to determine whether glycolysis-enhancing terazosin, alfuzosin, or doxazosin (TZ/AZ/DZ) is associated with lower hazard of developing dementia compared to tamsulosin. We identified 14,184 men with PD who had not taken TZ/DZ/AZ or tamsulosin previously. After matching for demographics, comorbidities, and disease duration, 1508 men (754 matched pairs) remained. This matching successfully reduced imbalance between the groups (Supplemental Table S1). After matching, the reduction was a statistically significant 25% reduction (HR = 0.75; 95% CI: 0.57, 0.98) in the hazard of being diagnosed with dementia (Fig. 4).

**Figure 4:**
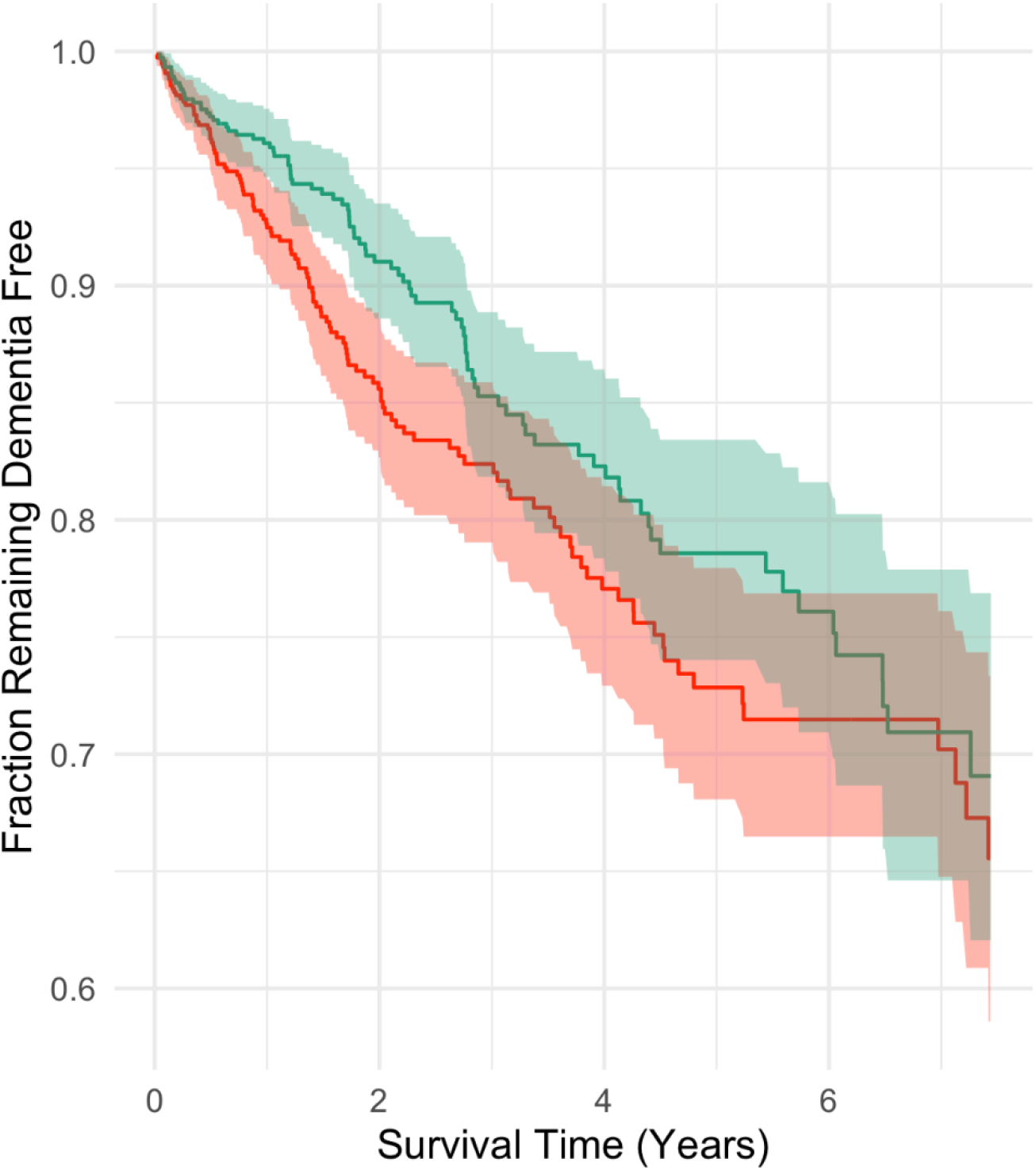
Fraction of Parkinson’s disease patients remaining dementia free in the Truven Database. Kaplan-Meier plot of 754 matched pairs of men aged 40 or older diagnosed with Parkinson’s disease (PD) and newly started on terazosin, alfuzosin, or doxazosin (green) or tamsulosin (red). The red and green lines denote cumulative incidence of PD patients remaining dementia free, and the shaded areas denote 95% CI.

## DISCUSSION

We tested the hypothesis that the glycolysis-enhancing drug terazosin is protective against cognitive dysfunction associated with PD. We found that VTA dopamine-depleted mice had increased interval timing variability, modeling cognitive deficits seen in PD patients. Rodents with VTA dopamine depletion and terazosin had preserved interval timing variability and a trend toward more VTA TH+ fluorescence compared to dopamine-depleted mice treated with vehicle. Finally, analysis of patient databases revealed that PD patients newly started on terazosin had less risk of developing dementia compared to PD patients taking tamsulosin, an α_1_-adrenergic receptor antagonist that does not impact glycolysis. Together these data provide evidence that terazosin is protective for cognitive, as well as motor, symptoms of PD. This is key as there are only a few minimally effective treatments for PD-related cognitive symptoms and no treatments that alter disease course.

Increased interval timing variability models some aspects of cognitive dysfunction in PD and Alzheimer’s disease patients (Gür et al., 2020; Kim et al., 2017; Larson et al., 2022; Parker et al., 2015; Singh et al., 2021). Importantly, the VTA is involved in PD and degenerates through the course of the disease (Alberico et al., 2015). Accordingly, we observed that VTA dopamine depletion increased interval timing variability, and terazosin reversed or prevented deficits. Furthermore, we found a non-statistically significant trend for more TH+ fluorescence in VTA dopamine-depleted mice treated with terazosin compared to those treated with DMSO alone. The level of VTA TH+ fluorescence correlates with interval timing variability (Gür et al., 2020). However, this correlation does not hold for VTA dopamine-depleted treated with terazosin as this treatment likely changed the relationship between VTA TH+ fluorescence and interval timing variability. Further, it is possible that terazosin might increase synaptic dopamine in the projection fields of VTA dopamine neurons, as was observed in nigrostriatal dopamine terminals (Cai et al., 2019). Our data establish that VTA dopamine depletion models PD-like increased interval timing variability, which is protected by terazosin.

From analysis of patient databases, we previously showed that terazosin is associated with fewer PD diagnostic codes linked with cognition, relative to tamsulosin (Cai et al., 2019). Here, we expand this finding and show that PD patients have less risk of developing dementia relative to patients taking tamsulosin. Because these data are derived from large administrative databases, it is difficult to discern whether these patients had interval timing deficits.

Our work has several limitations. First, there is no definitive rodent model of cognitive dysfunction in PD, in part because the underlying pathophysiology is complex and diverse. However, our model of VTA dopamine depletion models at least one characteristic of cognitive dysfunction in PD. Second, the mechanistic link between enhanced glycolysis and protection of dopamine neurons, dopamine release, or TH levels and/or production is unclear. However, it is possible that terazosin is protecting against VTA 6-OHDA-induced cell death or enhancing TH or dopamine similar to that observed in the nigrostriatal pathway (Cai et al., 2019). This is an important future direction to understand how glycolysis-enhancing therapeutics can modulate neurodegenerative disease. Third, our data from terazosin are retrospectively observational and depend on comparisons with tamsulosin (Gros et al., 2021; Sasane et al., 2021; Simmering et al., 2021).

These data converge with our prior work on motor symptoms of PD and provide evidence that terazosin is protective not only for neurons in the substantia nigra, but also in the ventral tegmental area (Alberico et al., 2015; Cai et al., 2019). Future studies will rigorously test this idea with a prospective and placebo-controlled randomized trial, in which motor and cognitive aspects of PD are carefully measured, and with cellular, molecular, and preclinical studies to understand how enhancing glycolysis modulates brain circuits.

## MATERIALS AND METHODS

### Mice

All experimental procedures were performed in accordance with the relevant guidelines of Protocol #0062039 and with the approval of the Institutional Animal Care and Use Committee (IACUC) at the University of Iowa. Wild-type male and female C57BL/6 mice were received from Jackson Labs (Bar Harbor, ME) at approximately 12–14 weeks of age and acclimated to the animal holding facility for 2 weeks. During acclimation, all mice were communally housed on a 12-hour light/dark cycle with *ad lib* access to laboratory rodent chow and water. To facilitate operant behavioral training (described below), mice were individually housed, weighed daily, and maintained on a restricted diet with *ad lib* access to water.

### Interval Timing Switch Task

The interval timing switch task is designed to capture an animal’s internal representation of time (Balci et al., 2008; Bruce et al., 2021; Larson et al., 2022; Tosun et al., 2016), as in other interval timing tasks (Emmons et al., 2020; Narayanan et al., 2012). Mice are trained to switch from a short to a long nosepoke after approximately 6 seconds. This switch is an explicit time-based decision that requires working memory for temporal rules and attention to the passage of time, and models cognitive deficits in neurodegenerative disease (Gür et al., 2020; Larson et al., 2022; Parker et al., 2015; Singh et al., 2021).

Briefly, mice were trained in standard operant chambers enclosed in sound-attenuated cabinets (MedAssociates, St. Albans, VT) that contained two light-equipped nosepoke response ports (left and right) on the front wall, a reward hopper located between the two nosepoke response ports, and another light-equipped nosepoke on the back wall, opposite the reward hopper. Operant training began by shaping the animal’s behavior. Trial initiation began with a response at the back nosepoke, at which point either the left or right front nosepoke was illuminated, and a response at the appropriate port resulted in a reward. Mice were then advanced to the interval timing switch protocol, in which each session was randomly organized into 50% short (6 seconds) and 50% long (18 seconds) trials. Either the left or right nosepoke was designated for short trials and the contralateral nosepoke for long trials (counterbalanced across experimental groups). A back nosepoke response initiated a trial, generating two identical light cues above the left and right nosepokes, along with an 8-kHz tone (72 dB) for both trial types. In short trials mice received a reward after 6 seconds for the first response at the designated nosepoke. In long trials, mice would begin by responding at the short-trial-designated nosepoke. When there was no reinforcement after 6 seconds, the mouse would switch to the long-trial-designated nosepoke until a reward was delivered after 18 seconds. Once mice were performing optimally, they were taken off food restriction and underwent stereotaxic surgical procedures (outlined below). Behavioral changes were assessed by retraining the mice in the same interval timing switch task. Only long switch trials were analyzed during both training periods.

### Surgical Procedures

On the day of surgery, 6-hydroxydopamine hydrobromide (6-OHDA; Millipore Sigma #162957, Darmstadt, Germany) and desipramine hydrochloride (Millipore Sigma #D3900) were freshly made and stored on ice away from light. 6-OHDA was prepared at 2 mg/ml in 0.03% ascorbic acid (AA) and desipramine at 4 mg/ml in 0.9% saline. Mice were anesthetized under 4.0% isoflurane at 400 ml/min, and surgical levels of isoflurane (1.5%–3.0%) were maintained at approximately 120 ml/min (SomnoSuite, Kent Scientific, Torrington, CT, USA). Desipramine (25 mg/kg) was injected intraperitoneally (IP) to protect norepinephrine terminals against 6-OHDA (Thiele et al., 2012; Torres & Dunnett, 2012). An incision was made along midline and bilateral craniotomies drilled above the VTA (AP -3.3, ML +/-1.1). Mice were randomly assigned to three different groups: 1) midbrain vehicle injections treated with dimethyl sulfoxide (DMSO; Vehicle DMSO); 2) VTA dopamine depletion treated with DMSO (6-OHDA DMSO); and 3) VTA dopamine depletion treated with terazosin (6-OHDA terazosin). The vehicle group received microinjections of 0.5 μl 0.03% AA bilaterally, and the dopamine-depletion groups received equal volumes of 6-OHDA prepared in AA (AP -3.3, ML +/-1.1, DV -4.6 at 10° laterally). Vehicle or 6-OHDA was infused over 10 minutes (0.05 µl/min; Legato 130 Syringe Pump, kd Scientific, Holliston, MA, USA), with a 5-minute wait period before removing the needle. After the incision was closed, mice were moved to a clean cage with *ad lib* access to food and either DMSO- or terazosin-treated water (described below). All mice recovered for one week before transitioning back to the interval timing switch task for approximately 3–4 weeks of post-surgical behavioral training.

### Mouse terazosin

Following surgery, terazosin (Tocris #1506, Minneapolis, MN) prepared in DMSO was delivered through *ad lib* access to treated water. Water for control mice was made with equivalent volumes of DMSO alone. Preparation of terazosin began with a 100-mM stock solution (42 mg/ml in DMSO) stored at -80 ° C. The stock was diluted 1:100 to make a 1-mM (0.42 μg/μl in water) working solution. We added 106 μL of working solution, equating to 45 μg terazosin (0.42 μg/μl x 106 μl), to 300 ml water, to which mice had free access. The concentration of the final drinking water was 0.15 μg/ml terazosin (45 μg/300 ml). An average mouse drinks approximately 5 ml of water per day (Nicolaus et al., 2016), so each mouse consumed ∼0.75 μg terazosin daily (0.15 μg/ml x 5 ml/day). Additionally, experimental mice weigh on average 0.025 kg (Nicolaus et al., 2016); thus, the terazosin dose received by our animals was ∼0.03 mg/kg/day (0.75 μg/day/0.025 kg). All terazosin- and DMSO-treated water was replaced every 2 days for the duration of the experiment post-surgery (Cai et al., 2019).

### Histology

Mice were anesthetized with ketamine (100 mg/kg IP) and xylazine (10 mg/kg IP) and transcardially perfused with cold phosphate-buffered saline (PBS) and 4% paraformaldehyde (PFA). Brains were removed and post-fixed in 4% PFA overnight, followed by immersion in 30% sucrose for approximately 48 hours. The fixed brains were sliced at 40-μm coronal sections of the midbrain, using a cryostat (Leica Biosystems, Deer Park, IL). Sections were then blocked for one hour in 5% normal goat serum (NGS) in PBST (0.3% Triton X-100 in 1x PBS). After blocking, sections were incubated overnight at 4 °C in rabbit anti-tyrosine hydroxylase polyclonal antibody (Abcam #ab6211, Cambridge, UK) diluted to 1:1000 in 5% NGS. The sections were washed with PBST three times over 30 minutes before a 2-hour incubation in goat anti-rabbit IgG (H+L) Alexa Fluor 568 secondary antibody (Invitrogen #A-11036, Waltham, MA) diluted to 1:1000 in 5% NGS. After three more PBST washes, the sections were mounted with ProLong Diamond Antifade Mountant with DAPI (Invitrogen #P36962) on Superfrost microscope slides (Fisher Scientific, Waltham, MA).

Brain sections were imaged using an Olympus VS120 microscope (Olympus, Center Valley, PA). Histological targeting of the VTA was confirmed by two independent authors, one blinded to treatment conditions. Eight mice, in which the injection missed the VTA, were excluded. For the remaining mice, the Count and Measure analysis tool in CellSens Dimension Desktop (Olympus, Shinjuku, Tokyo, Japan) was used to quantify tyrosine hydroxylase (TH) fluorescence levels in the VTA. Fluorescence levels in each animal were determined by averaging the mean pixel intensities of three VTA sections. Anterior-posterior coordinates of the VTA were determined following prior literature and ranged from -3.1 to -3.5, respectively. Outlines of the regions of interest were referenced from Franklin & Paxinos, 2008.

### Statistics

We analyzed effects of dopamine depletion and terazosin using a generalized linear model (*lm* in R). Post-hoc comparisons were performed using estimated marginal means (*emmeans* in R) and Tukey’s correction for multiple comparisons. Four-to-five sessions of interval timing behavior were collected and analyzed per mouse, both before surgery and approximately 16 days post-surgery. Switch time coefficients of variability (CV) and mean switch times were normalized to each mouse’s pre-surgical baseline to account for animal-specific variability in timing behavior. All statistics was reviewed by the Biomedical Epidemiology Research and Design core in the Institute for Clinical and Translational Sciences at the University of Iowa.

### Administrative Database Search

Using the IBM Marketscan Commercial Claims and Encounters and the Medicare Coordination of Benefits databases of health insurance claims, we identified men aged 40 or older taking terazosin, alfuzosin, or doxazosin (collectively, TZ/DZ/AZ) or tamsulosin, not in conjunction with finasteride or dutasteride. Men who switched between the TZ/DZ/AZ and tamsulosin classes were excluded. To ensure that we identified men who were newly started on TZ/DZ/AZ or Tamsulosin, we required: 1) at least 12 months of enrollment prior to the observed first dispensing date with prescription drug coverage; 2) at least two dispensing events to occur in the first year following the first dispensing date; 3) the PD diagnosis date must have occurred before the TZ/DZ/AZ or tamsulosin start date; and 4) the men must have been free of a dementia diagnosis (ICD-9-CM: 289.9, 290.0, 290.1, 290.2, 290.3, 290.4, 290.43, 294.1, 294.8, 331.0, 331.1, 348.3 or ICD-10-CM: F01.51, F03.90, F05, F06.0, F06.8, F29) at start of medication. To ensure that we identified new cases of PD, the first observed diagnosis of PD or dispensing of levodopa must have occurred within at least 12 months of TZ/DZ/AZ or tamsulosin start date; health insurance claims data do not include detailed measures of PD severity, e.g., Unified Parkinson’s Disease Rating Scale scores or cognitive function. The administrative database search and subsequent analysis described below was performed on a fully deidentified secondary database and was therefore exempt from institutional review board approval.

### Administrative Database Analysis

Men taking TZ/DZ/AZ were matched to men taking tamsulosin in a two-stage process. First, we estimated a propensity score model incorporating the following criteria: age; health care utilization during the lookback period (rate of hospitalization, rate of outpatient encounters); baseline health status (mean number of diagnoses per outpatient encounter, rate of unique outpatient diagnoses, the 29 Elixhauser comorbidities); factors for prescribing decision (diagnosis of benign prostatic hyperplasia, diagnosis of slow urinary stream, diagnosis of abnormal prostate-specific antigen (PSA), diagnosis of orthostatic hypotension, diagnosis of other hypotension, procedural claim for testing PSA, procedural claim for a uroflow study, procedural claim for a cystometrogram); and medication start date. The propensity score model was estimated using a boosted tree with the depth and number of rounds selected by testing performance on a 25% held-out validation set. Once the depth and number of rounds were selected, we used the entire data set to generate and estimate the propensity score model. Second, we required the time between the diagnosis of PD and the medication start date to be +/- 180 days between possible matches. This was done to ensure a similar duration of disease and to potentially reduce unobserved heterogeneity due to differing PD severity. Within the set of possible matches based on the time between PD diagnosis and the medication start date, we selected the nearest possible match based on the log odds estimated by our propensity score model. We imposed a 20% pooled standard deviation caliper to exclude poor matches. After matching, we followed the medical records of men for 8 years to track the rates of developing dementia. We compared the hazard of developing dementia with Kaplan Meier survival curves and Cox proportional hazards regression. Standard errors were clustered to account for the propensity score matching.

## Data Availability

All data relevant to the human Truven database produced in the present study are available upon reasonable request to the authors. All animal data produced are available online at: https://narayanan.lab.uiowa.edu/article/datasets

## ACKNOWLEDGEMENTS

The Truven database was provided by the University of Iowa. This work was supported by a faculty fellowship through the Iowa Neuroscience Institute to JES and NIH R01s MH116043, NS120987 to NSN.

## DATA AVAILABILITY

All code and raw data are available at https://narayanan.lab.uiowa.edu.

## AUTHOR CONTRIBUTIONS

MAW, KS, and NSN designed the animal experiments. JES and NSN designed the human PD database analysis. MAW, KS, and MO performed all animal experiments. MAW and KS independently verified histological targeting. EET, KS, and GMA performed histological immunofluorescent analysis. MAW, KS, and QZ maintained and delivered terazosin. MAW, KS, JES, and NSN performed all statistical analyses. MAW, KS, JES and NSN wrote the manuscript, and all authors reviewed and revised the manuscript.

## Supplemental

**Table S1:**
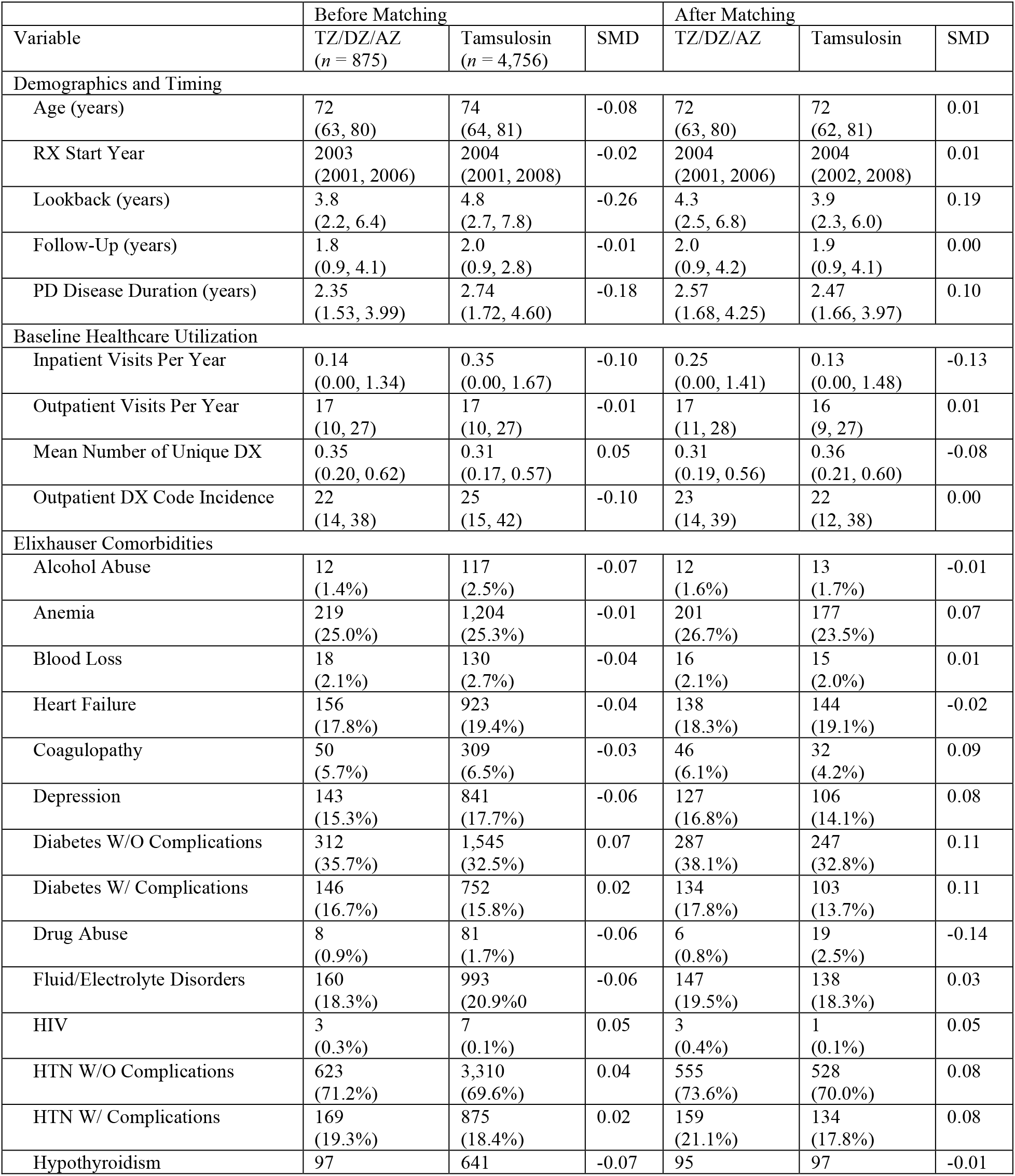
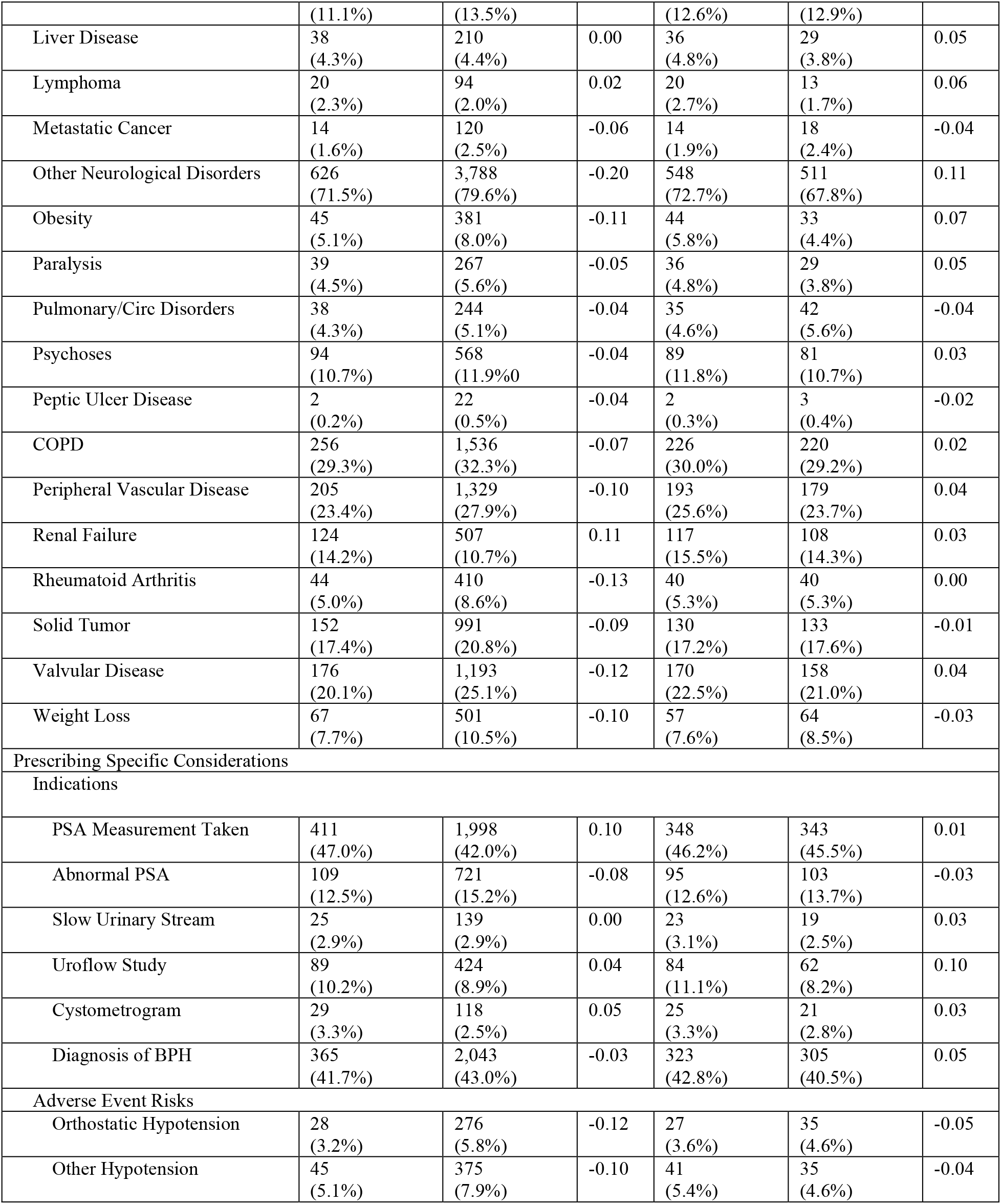
Cohort Summary Measures Before and After Matching. Values reported are median and inner quartile range for continuous variables or count and percent for binary variables. SMD is the standardized mean difference or Cohen’s *d*, a measure of effect size. Balance was obtained (absolute value of SMD <= 0.1) on nearly all measures included in the propensity score model or the disease duration.

